# SARS-CoV-2 Infection and Long-Term Risk of Cardiovascular and Renal Morbidity

**DOI:** 10.1101/2025.01.14.25320050

**Authors:** Heather A. Boyd, Thor Grønborg Junker, Tor Biering-Sørensen, J Jan Wohlfahrt, Anders Hviid

## Abstract

**Importance:** Cardiovascular and renal consequences of severe acute respiratory syndrome coronavirus 2 (SARS-CoV-2) infection have been studied intensively in high-risk groups, but the consequences of mild infection for the general population, particularly beyond the acute phase of infection, remain unclear.

**Objective:** To examine long-term associations between SARS-CoV-2 infection and cardiovascular and kidney disease (CVD, KD) in the general population, with emphasis on age and vaccination status at the time of infection, mild infection, and SARS-CoV-2 variant.

**Design:** Register-based cohort study.

**Setting:** Denmark

**Participants:** All Danish residents with ≥1 PCR test for SARS-CoV-2 infection, March 2020-December 2022.

**Exposure:** Positive PCR test for SARS-CoV-2 infection.

**Main Outcomes:** Hazard ratios for 15 CVD outcomes and six KD outcomes, comparing persons testing positive for SARS-CoV-2 infection and persons who only ever tested negative for infection.

**Results:** The cohort for CVD analyses included 4,508,489 persons without pre-existing CVD (median follow-up 25.2 months/person, interquartile range [IQR] 21.7-27.5 months); 2,698,261 persons (59.8%) tested positive for SARS-CoV-2 infection during the study period. The cohort for the KD analyses included 5,150,480 persons without pre-existing KD (median follow-up 25.1 months, IQR 21.7-27.4 months), 2,983,233 (57.9%) of whom tested positive for infection. SARS-CoV-2 infection was associated with slight increases in the risks of pulmonary embolism, venous embolism/thrombosis, arrhythmias, chronic renal failure, unspecified renal failure, and other/unspecified KD up to a year after infection; infection was not associated with the other disease groups tested. The strongest associations between SARS-CoV-2 infection and CVD and KD were observed among unvaccinated persons and persons infected with earlier (pre-omicron) variants.

**Conclusions and Relevance:** We found little evidence that infection with SARS-CoV-2 was associated with increased long-term risks of CVD or KD in the general population. Increased CVD risks associated with SARS-CoV-2 infection appeared limited to three outcomes (pulmonary embolism, venous embolism/thrombosis, arrhythmias) and the potential increases in risk were small. Our KD results also suggested that any persistent risks associated with SARS-CoV-2 infection were minimal; however, these findings need to be confirmed in other populations. Most importantly, in a largely vaccinated population, long-term CVD and KD risks differed little for omicron-infected and uninfected persons.

**KEY POINTS:** 

**Question:** Does SARS-CoV-2 infection increase the long-term risks of cardiovascular disease (CVD) or kidney disease (KD) in a general population cohort with predominantly mild infection?

**Findings:** In a cohort study of >4.5 million persons that compared SARS-CoV-2 test-positive and test-negative persons, relative risks were slightly increased for 6 of 21 CVD and KD outcomes up to 1 year after infection. Among vaccinated individuals and omicron-infected persons, existing associations diminished dramatically.

**Meaning:** In the general population, long-term increases in CKD and KD risks associated with infection were small, limited to a few outcomes, and attenuated with vaccination and the omicron variant.

---

Although the severe acute respiratory syndrome coronavirus 2 (SARS-CoV-2) pandemic has subsided, the long-term health consequences of past SARS-CoV-2 infection remain relevant. Global endemicity and continuous emergence of variants with epidemic potential make it crucial to identify downstream morbidity robustly associated with SARS-CoV-2 infection, pinpoint high-risk groups, and assess whether the risks are mitigated by vaccination.

In humans, SARS-CoV-2 binds to the cellular receptor form of angiotensin-converting enzyme 2 (ACE2) in the lungs.^1^ ACE2 is also highly expressed in the heart and kidneys, and severe cardiovascular and renal morbidity associated with acute SARS-CoV-2 infection is well-documented.^2,3^ Cardiovascular and renal morbidity after SARS-CoV-2 infection has also been extensively studied in high-risk subgroups (e.g. persons hospitalized for severe coronavirus disease 2019 [COVID-19], persons with long COVID-19 syndrome).^4-6^ However, potential links between uncomplicated SARS-CoV-2 infection in the general population and morbidity after resolution of acute infection are less well-elucidated. In some studies, SARS-CoV-2 infection was associated with an increased long-term risk of cardiovascular disease (CVD);^7,8^ others found that although the short-term risk of CVD increased dramatically, it declined substantially one month post-infection.^4,9,10^ Long-term renal outcomes following SARS-CoV-2 infection have received even less attention.^8,11,12^

Discrepancies in reported long-term risks of CVD after SARS-CoV-2 infection are likely due to differences in definition of infection, comparison cohort, inclusion/exclusion criteria, and ability to handle changing SARS-CoV-2 infection status, potential confounders, and vaccination status. To address previous methodological issues and provide new insight into the long-term consequences of SARS-CoV-2 infection, we conducted a cohort study using data from Denmark’s universally-available, nationwide SARS-CoV-2 infection testing system to examine associations between SARS-CoV-2 infection and CVD and kidney disease (KD) in the general population up to 33 months after infection, with emphasis on age and vaccination status at the time of infection, mild infections, and SARS-CoV-2 variant.

## METHODS

### Study Cohorts and Follow-Up

Using information from Denmark’s SARS-CoV-2 infection testing system registered in the Danish Microbiology Database,^13^ we constructed a study population that included all Danish residents with at least one PCR test for SARS-CoV-2 infection in the period 1 March 2020 to 31 December 2022. We excluded persons registered in the National Patient Register^14^ with chromosomal abnormalities or congenital syndromes with multiple organ system defects before the start of follow-up (Online Supplement, Definitions).

The cohort used to evaluate associations with CVDs further excluded persons with CVD, congenital heart defects, or other relevant heart-related diagnoses registered before the start of follow-up, while the cohort used to address KD excluded persons with KD, congenital kidney defects, and other relevant kidney-related diagnoses (Online Supplement, Definitions). For each cohort, follow-up began 30 days after a person’s first PCR test for SARS-CoV-2 in the study period and continued until the first of the following: 1) first registered CVD or KD diagnosis, depending on the outcome of interest; 2) death; 3) emigration; 4) registered as “missing” in the Civil Registration System;^15^ 5) 31 December 2022. Use of the 30-day washout period diminished potential surveillance bias produced by mandatory SARS-CoV-2 testing of all in- and outpatients during the pandemic.

### SARS-CoV-2 Infection (Exposure)

The exposure of interest was PCR-confirmed SARS-CoV-2 infection status, treated as a time-dependent variable; a person whose first test in the study period was negative contributed time to the uninfected group until they received a later positive test (if ever) (Supplementary Figure 1). Information on test dates and results (positive or negative for SARS-CoV-2) were obtained from the Danish Microbiology Database. A presumptive SARS-CoV-2 variant was assigned based on the predominant variant circulating at the time of PCR testing.^16^ We defined severe infection as an infection confirmed ≤14 days before or ≤2 days after a hospital admission or outpatient contact lasting at least 12 hours and accompanied by a COVID-19-related International Classification of Diseases, version 10 (ICD-10), diagnosis code in the National Patient Register.

### Outcomes

We identified persons with incident CVD and KD during follow-up based on registration of ICD-10 codes in the National Patient Register. To avoid making assumptions about which conditions were likely to be associated with SARS-CoV-2 infection, we considered a broad range of outcomes, including 15 groups of CVDs and 7 KD groups (Online Supplement, Definitions).

### Covariates

Information on sex and date of birth was obtained from the Civil Registration System. We obtained information on the comorbidities included in the updated Charlson Comorbidity Index (uCCI^17^) from the National Patient Register and estimated degree of comorbidity at the start of follow-up (none, uCCI=0; mild/moderate, uCCI=1-2; severe, uCCI≥3). We obtained information on vaccination status from the Danish Vaccination Register^18^ and classified persons as having received 0, 1, 2 or ≥3 doses of SARS-CoV-2 vaccine (time-dependent variable).

### Statistical analysis

Using Cox regression with age as the underlying time scale,^19^ we estimated hazard ratios comparing rates of CVD and KD in persons with PCR-confirmed SARS-CoV-2 infection with those in persons with only negative SARS-CoV-2 test results. Because the proportional hazards assumption was not fulfilled across the entire study period, our main analyses present hazard ratios for five intervals from the start of follow-up, within which the baseline hazards were piecewise constant: 1) Day 1; 2) Day 2 to <1 month; 3) 1-5 months; 4) 6-11 months; 5) ≥12 months. Persons could not contribute outcomes to multiple outcome groups; we considered only the first CVD and the first KD diagnosis registered for each person. All estimates were adjusted for sex, calendar period, and degree of comorbidity at the start of follow-up.

In additional analyses, we investigated how the observed associations changed when we restricted to non-severe infections and stratified by age, SARS-CoV-2 variant, and vaccination status. Because the results of the main analyses revealed strong associations between infection and many outcomes in the first month of follow-up (consistent with previously-documented associations with acute infection and possible residual surveillance bias), and our focus was on associations with later morbidity, these analyses concentrated on the period beginning 1 month after the start of follow-up. Due to small numbers of outcomes in some groups, these analyses were not stratified by time since the start of follow-up.

Widespread PCR testing in Denmark ended on 10 March 2022, when the National Board of Health recommended that only high-risk persons with COVID-19 symptoms be tested.^20^ To ensure that our results were not biased towards the null by misclassification as uninfected of untested persons infected after this date, we conducted a sensitivity analysis where follow-up ended on 10 March 2022.

## RESULTS

### Cohorts

In the CVD analyses, we followed 4,508,489 persons for 8,909,627 person-years (median, 25.2 months, interquartile range [IQR] 21.7-27.5 months), during which 2,698,261 persons tested positive for SARS-CoV-2 infection. Supplementary Figure 2 shows the distribution of follow-up time in the CVD cohort across the study period, by exposure category. In the KD analyses, 5,150,480 persons were followed for 10,146,408 person-years (median 25.1 months, IQR 21.7-27.4 months), with 2,983,233 persons testing positive for infection. The distribution of follow-up time in the KD cohort across the study period was similar to that in the CVD cohort (data not shown). Table 1 shows the distribution of age (CVD cohort: median 34.2 years, IQR 18.5-52.3 years; KD cohort: median 39.0 years, IQR 20.8-57.5 years), sex, and comorbidities in the cohorts at the start of follow-up, by first SARS-CoV-2 PCR test result in the study period.

**Table 1.**
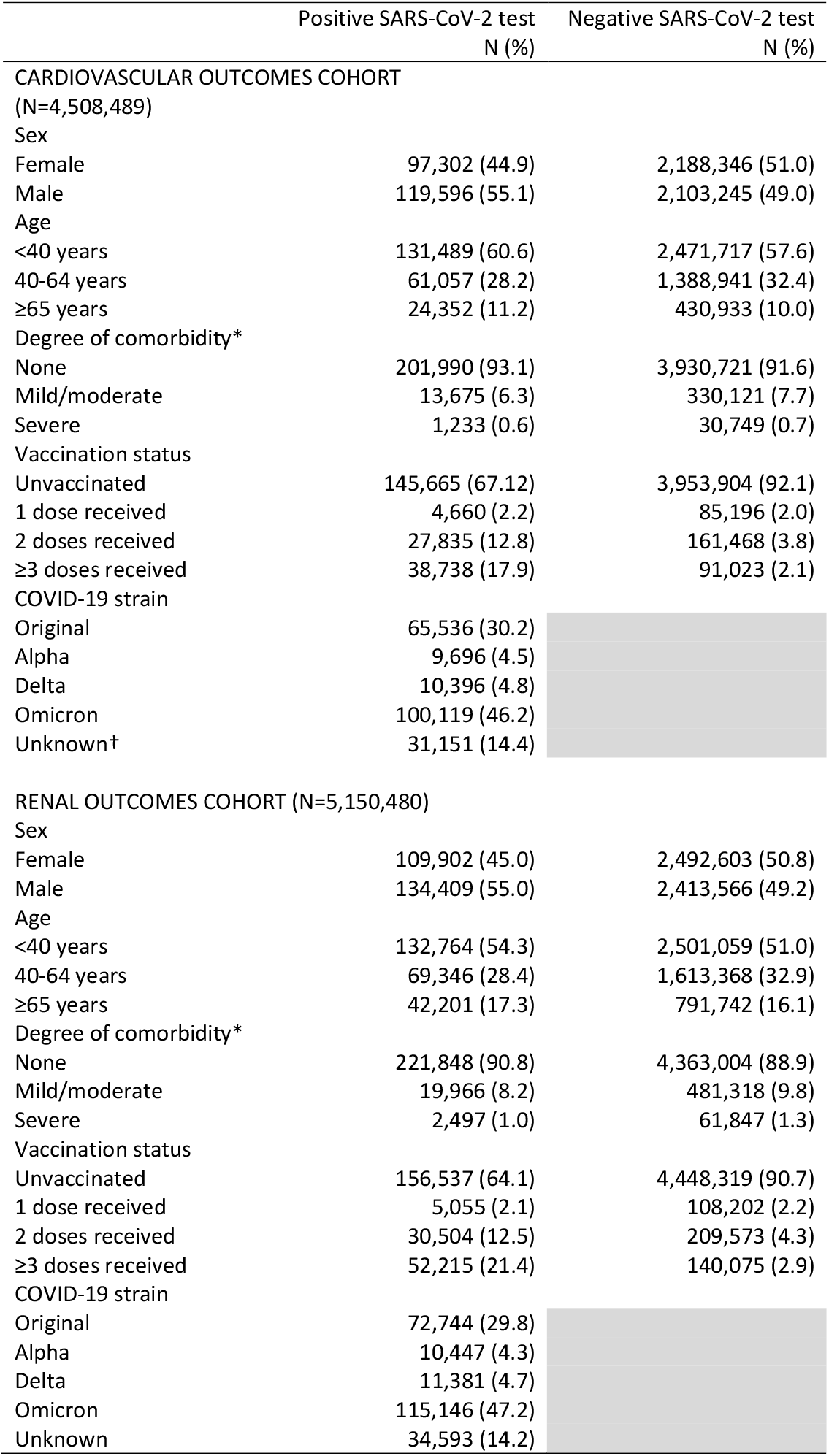
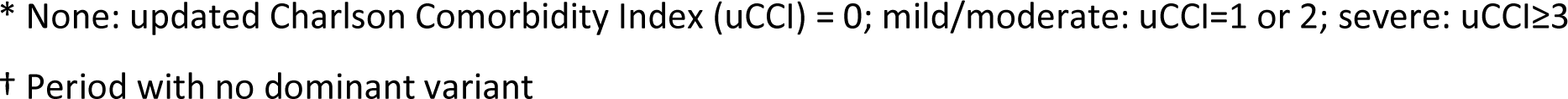
Characteristics of persons included in the two study cohorts at their first SARS-CoV-2 test in the study period, by test outcome, Denmark, 2020-2022.

|  | Positive SARS-CoV-2 test<br>N (%) | Negative SARS-CoV-2 test<br>N (%) |
| --- | --- | --- |
| CARDIOVASCULAR OUTCOMES COHORT |  |  |
| (N=4,508,489) |  |  |
| Sex |  |  |
| Female | 97,302 (44.9) | 2,188,346 (51.0) |
| Male | 119,596 (55.1) | 2,103,245 (49.0) |
| Age |  |  |
| <40 years | 131,489 (60.6) | 2,471,717 (57.6) |
| 40-64 years | 61,057 (28.2) | 1,388,941 (32.4) |
| ≥65 years | 24,352 (11.2) | 430,933 (10.0) |
| Degree of comorbidity* |  |  |
| None | 201,990 (93.1) | 3,930,721 (91.6) |
| Mild/moderate | 13,675 (6.3) | 330,121 (7.7) |
| Severe | 1,233 (0.6) | 30,749 (0.7) |
| Vaccination status |  |  |
| Unvaccinated | 145,665 (67.12) | 3,953,904 (92.1) |
| 1 dose received | 4,660 (2.2) | 85,196 (2.0) |
| 2 doses received | 27,835 (12.8) | 161,468 (3.8) |
| ≥3 doses received | 38,738 (17.9) | 91,023 (2.1) |
| COVID-19 strain |  |  |
| Original | 65,536 (30.2) |  |
| Alpha | 9,696 (4.5) |  |
| Delta | 10,396 (4.8) |  |
| Omicron | 100,119 (46.2) |  |
| Unknown† | 31,151 (14.4) |  |
| RENAL OUTCOMES COHORT (N=5,150,480) |  |  |
| Sex |  |  |
| Female | 109,902 (45.0) | 2,492,603 (50.8) |
| Male | 134,409 (55.0) | 2,413,566 (49.2) |
| Age |  |  |
| <40 years | 132,764 (54.3) | 2,501,059 (51.0) |
| 40-64 years | 69,346 (28.4) | 1,613,368 (32.9) |
| ≥65 years | 42,201 (17.3) | 791,742 (16.1) |
| Degree of comorbidity* |  |  |
| None | 221,848 (90.8) | 4,363,004 (88.9) |
| Mild/moderate | 19,966 (8.2) | 481,318 (9.8) |
| Severe | 2,497 (1.0) | 61,847 (1.3) |
| Vaccination status |  |  |
| Unvaccinated | 156,537 (64.1) | 4,448,319 (90.7) |
| 1 dose received | 5,055 (2.1) | 108,202 (2.2) |
| 2 doses received | 30,504 (12.5) | 209,573 (4.3) |
| ≥3 doses received | 52,215 (21.4) | 140,075 (2.9) |
| COVID-19 strain |  |  |
| Original | 72,744 (29.8) |  |
| Alpha | 10,447 (4.3) |  |
| Delta | 11,381 (4.7) |  |
| Omicron | 115,146 (47.2) |  |
| Unknown | 34,593 (14.2) |  |

\* None: updated Charlson Comorbidity Index (uCCI) = 0; mild/moderate: uCCI=1 or 2; severe: uCCI ≥ 3
† Period with no dominant variant

### Main Results: Cardiovascular Disease

During the follow-up period, 70,885 persons were diagnosed with CVD, of whom 16,475 had previously tested positive for SARS-CoV-2. Of these, 1,767 developed CVD within 30 days of the start of follow-up, 6,783 did so within 1-5 months, and 6,324 developed CVD after 6-11 months. Among persons with a positive SARS-CoV-2 test, arrhythmias were the most frequently registered diagnosis (N=4,538, 27.5%), followed by ischemic heart disease (N=2,327, 14.1%), venous embolism/thrombosis (N=1,820, 11.0%), and cerebral infarction (N=1,564, 9.5%) (Supplementary Table 1).

Figure 1 (Supplementary Table 1) shows hazard ratios for 15 CVD groups comparing persons with a positive SARS-CoV-2 test and persons with only negative test results, by length of follow-up after the first positive test in the study period. Hazard ratio magnitudes on Day 1 ranged from 2.51 to 16.1. Thereafter, the rates of most CVDs did not differ for persons with and without a positive SARS-CoV-2 test, regardless of length of follow-up. Exceptions included pulmonary embolism, venous embolism/thrombosis, and arrhythmias, the rates of which were increased in test-positive persons within 30 days of starting follow-up (hazard ratio [HR] 4.14, 95% confidence interval [CI] 3.57-4.79; HR 1.42, 95% CI 1.22-1.66; and HR 1.19, 95% CI 1.07-1.32, respectively). Rates of pulmonary embolism remained somewhat elevated for 6 months (HR 1.21, 95% CI 1.08-1.36) in persons with a positive test, before dropping to levels observed in never-infected persons (Figure 1, Supplementary Table 1), while rates of venous embolism/thrombosis and arrhythmias remained slightly elevated for up to a year (HR range 1.05-1.12) (Figure 1, Supplementary Table 1).

**Figure 1.**
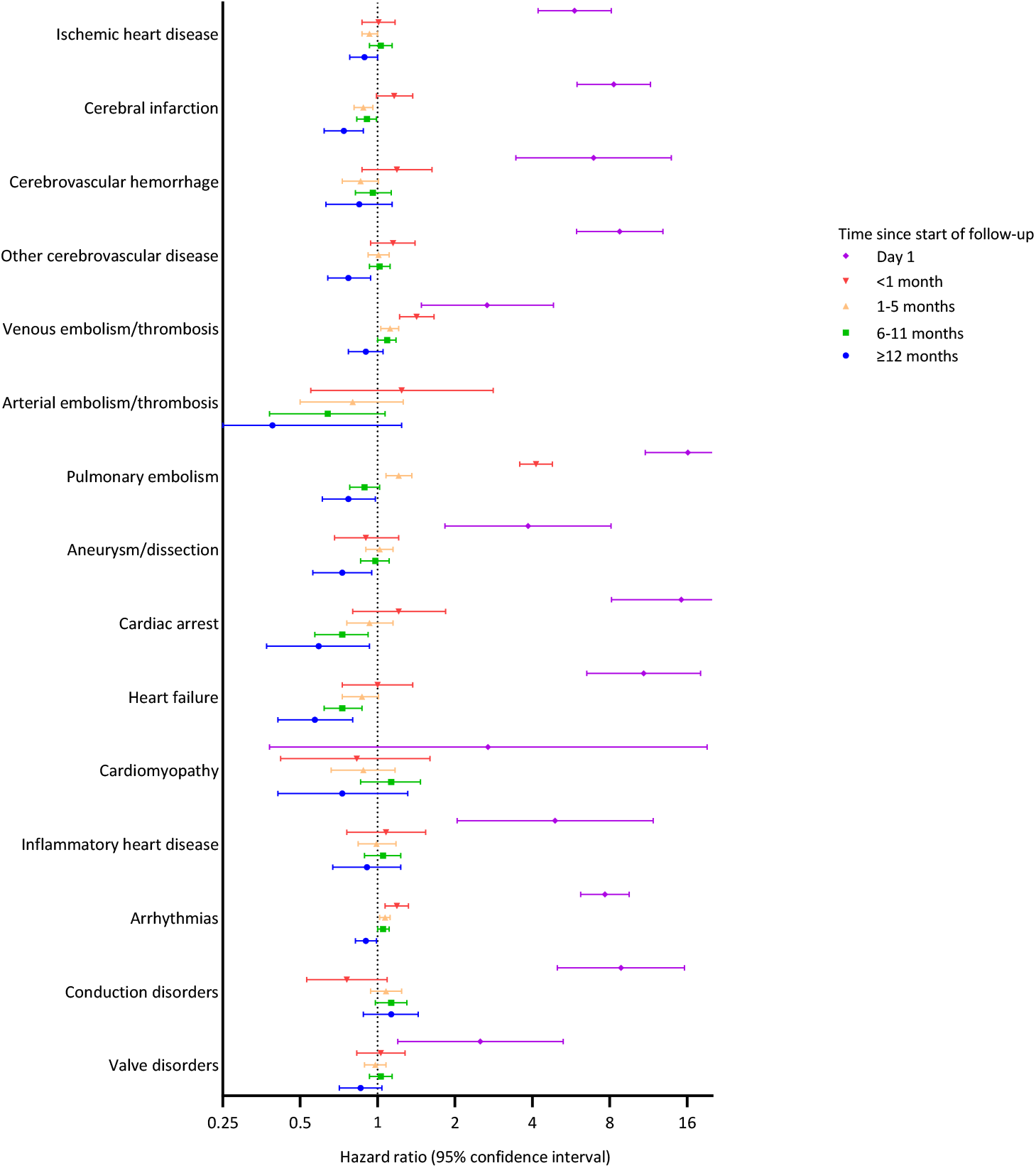
Hazard ratios for 15 cardiovascular disease groups comparing persons with a positive SARS-CoV-2 test and persons with only negative SARS-CoV-2 test results, by length of follow-up after the first positive test in the study period, Denmark, March 2020-December 2022. Purple: 1 day of follow-up. Red: >1 day but <1 month of follow-up. Orange: 1-5 complete months of follow-up. Green: 6-11 complete months of follow-up. Blue: 12 or more months of follow-up.

### Main Results: Kidney Disease

Of 16,016 persons diagnosed with KD during follow-up, 3,897 had previously tested positive for SARS-CoV-2 infection. Of these, 518 developed KD within 30 days of the start of follow-up, 1,393 did so within 1-5 months, and 1,547 did so after 6-11 months. Among persons testing positive for SARS-CoV-2, acute kidney injury (AKI) and other acute KDs were the most frequently registered diagnoses (N=2,017, 51.8%), followed by other/unspecified KD (N=1,173, 30.1%) and chronic renal failure (N=481, 12.3%) (Supplementary Table 2).

Figure 2 (Supplementary Table 2) shows hazard ratios for six KD groups, by length of follow-up after the first positive test in the study period. On Day 1, rates of KD were four to 28 times higher for persons testing positive for SARS-CoV-2 than for persons testing negative. Association magnitudes diminished dramatically thereafter, reaching a nadir after 1-5 months. Hazard ratios appeared to increase again in the following 6 months for all outcomes except AKI, although not all associations were statistically significant (Figure 2, Supplementary Table 2).

**Figure 2.**
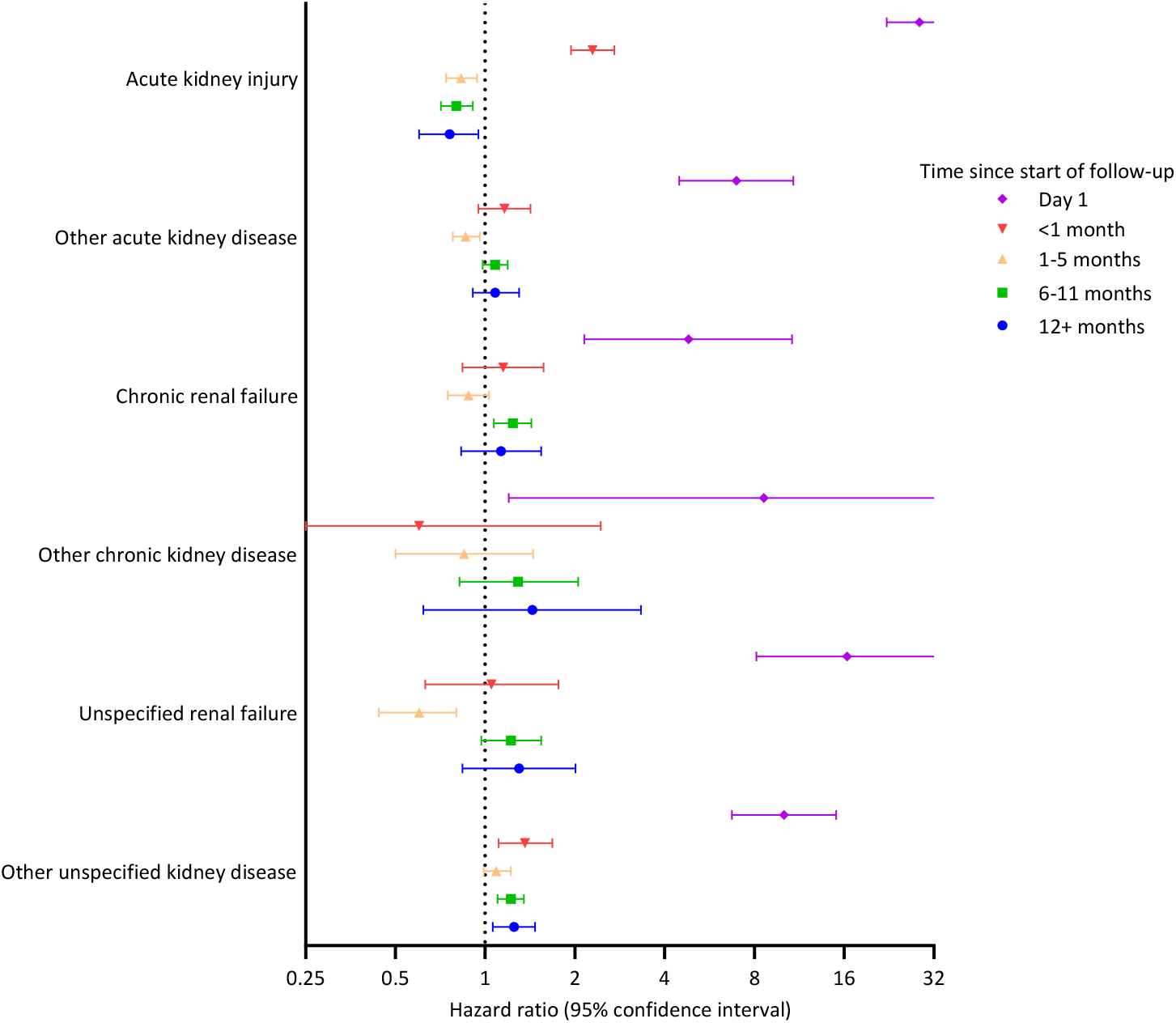
Hazard ratios for 6 kidney disease groups comparing persons with a positive SARS-CoV-2 test and persons with only negative SARS-CoV-2 test results, by length of follow-up after the first positive test in the study period, Denmark, March 2020-December 2022. Purple: 1 day of follow-up. Red: >1 day but <1 month of follow-up. Orange: 1-5 complete months of follow-up. Green: 6-11 complete months of follow-up. Blue: 12 or more months of follow-up.

### Age, Infection Severity, SARS-CoV-2 Variant, and Vaccination Status

Associations were generally consistent across age groups (Supplementary Figure 3, Supplementary Table 3). Possible exceptions included the associations with pulmonary embolism, chronic renal failure, and unspecified renal failure, which were strongest in persons <40 years of age (HR 1.25, 95% CI 0.98-1.59; HR 3.13, 95% CI 1.93-5.10; and HR 3.07, 95% CI 1.47-6.40, respectively) and decreased or disappeared with increasing age, and other/unspecified KD, which was strongest among persons aged ≥65 years (HR 1.34, 95% CI 1.13-1.59).

When we compared persons with non-severe infection and persons with only negative test results, there was little evidence of an association between SARS-CoV-2 infection and most outcomes (Supplementary Table 4). Non-severe infection was associated with minor increases in the risk of venous embolism/thrombosis (HR 1.09, 95% CI 1.02-1.16), arrhythmias (HR 1.05, 95% CI 1.01-1.09), conduction disorders (HR 1.09, 95% CI 0.98-1.21), and other/unspecified KD (HR 1.13, 95% CI 1.04-1.22).

Where associations existed, infection with the original SARS-CoV-2 strain and the alpha and delta variants were associated with greater increases in CVD rates than the omicron variant (Supplementary Figure 4, Supplementary Table 5). The omicron variant was only associated with increased rates of arrhythmias, venous embolism/thrombosis, and conduction disorders, and then only weakly (HR range: 1.04-1.18). Otherwise, there was no consistent trend in strength of association according to the remaining variant types. For KD, SARS-CoV-2 variant analyses were largely inconclusive owing to wide confidence intervals (Supplementary Figure 4, Supplementary Table 5). However, the original strain and the alpha and delta variants were associated with modestly increased risks of other/unspecified KD, whereas there was no association with the omicron variant.

Among persons who received two doses of SARS-CoV-2 vaccine, SARS-CoV-2 infection was only associated with increased risks of arrhythmias, chronic renal failure, and possibly other/unspecified KD (HR 1.11, 95% CI 1.00-1.23; HR 2.48, 95% CI 1.76-3.49; and HR 1.15, 95% CI 0.95-1.39, respectively; Supplementary Figure 5, Supplementary Table 6). Among persons who received three or more vaccine doses, there was little evidence of association between SARS-CoV-2 infection and any CVD or KD, with the possible exceptions of venous embolism/thromboembolism (HR 1.10, 95% CI 0.96-1.25) and conduction disorders (HR 1.20, 95% CI 0.95-1.50).

### Sensitivity Analyses

Ending follow-up in March 2022 attenuated some estimates and decreased the stability of the results due to reduced numbers of events, but did not change our conclusions (Supplementary Tables 7 and 8).

## DISCUSSION

### Key Findings

In a study that included most of the Danish population in one or both study cohorts, we found little evidence that SARS-CoV-2 infection was associated with increased risk of CVD or KD beyond the first day of follow-up, with a few exceptions. Our results suggested that SARS-CoV-2 infection might be associated with slight increases in the risks of pulmonary embolism, venous embolism/thrombosis, arrhythmias, chronic renal failure, unspecified renal failure, and other/unspecified KD up to a year after SARS-CoV-2 infection. The strongest associations were observed among unvaccinated persons and persons infected with earlier (pre-omicron) variants.

### Comparison with Previous Study Findings

Contrary to previous studies,^4,9,10^ apart from associations observed on the first day of follow-up, we found only limited evidence of associations between SARS-CoV-2 infection and CVD in the first 30 days of follow-up, and the associations we did observe were weak. Surveillance bias in previous studies and differences in study populations likely explain much of this difference in study findings. In most countries during the pandemic, all hospital in- and outpatients were tested for SARS-CoV-2 infection at first contact. Persons seeking care for, or at high risk of, CVD were therefore more likely to be tested and found to be infected with SARS-CoV-2 than were persons without CVD, artificially inflating hazard ratios early in the follow-up period (i.e. surveillance bias, the residual effects of which likely account for our Day 1 results). Whereas ours was a general population cohort with predominantly mild or asymptomatic infection, other study populations were hospital-based^10^ or oversampled persons with severe infection and therefore potentially more comorbidities.^9^

Consistent with the aforementioned studies,^4,9,10^ we found little evidence of associations between SARS-CoV-2 infection and CVD beyond the first 30 days of follow-up, with the exception of slight increases in the risks of pulmonary embolism in the first 6 months and venous embolism/thromboembolism and arrhythmias in the first year. Our CVD results were also consistent with recent findings that vaccination greatly reduced the risk of CVD events after SARS-CoV-2 infection.^21^

In contrast, two previous studies reported strong associations between SARS-CoV-2 infection and many CVDs more than 30 days post-infection.^7,8^ However, both studies had design features that may have inflated CVD rates among infected persons. In one study,^7^ infected persons were identified through SARS-CoV-2 testing required for contact with the American Veterans Health Administration (VA) system, whereas the comparison groups were either historical (pre-pandemic) or contemporary but not necessarily tested for SARS-CoV-2 by the VA. SARS-CoV-2-infected persons had more contact with the VA system during follow-up and were therefore more likely to be diagnosed with CVD. Similarly, a binational meta-analysis compared persons with a positive SARS-CoV-2 test or a COVID-19 diagnosis with persons with no record of a positive test or diagnosis, rather than with persons with a negative test, while considering only hospital outcomes in one of the countries.^8^ Nevertheless, despite significant between-study differences in design and potential accompanying biases, all studies to date have suggested that some increase in the risks of pulmonary embolism, venous embolism/thrombosis, and arrhythmias may persist for up to a year after SARS-CoV-2 infection.

We confirmed the known strong association with AKI shortly after infection;^22^ we also found a modest association between SARS-CoV-2 infection and other/unspecified KD in the first 30 days of follow-up. Few studies have examined long-term associations between SARS-CoV-2 infection and KD, particularly in general population cohorts not hospitalized for infection. One study found that the risk of AKI remained doubled in the post-acute period, with the risk of end-stage renal disease also increased.^8^ Another reported increased risks of chronic KD in the post-acute period, with the strongest associations in persons ≥60 years of age,^11^ whereas we reported a trebling of the risk of chronic renal failure among infected persons <40 years of age and no association in persons ≥65 years.

### Strengths and Potential Limitations

Our study’s strengths included its large size, long follow-up period, and comparison of persons with PCR-confirmed SARS-CoV-2 infection with persons who were PCR-tested and found not to be infected (rather than presuming non-infection in untested persons). Inclusion of Denmark’s entire population, use of data from a free, widely-used, national infection testing program spanning the pandemic period,^23^ and inclusion of all age groups and the entire spectrum of infection phenotypes from asymptomatic to severe, minimized the risk of selection bias. Obtaining data on exposure and outcomes from national registers with prospective registration eliminated the possibility of recall bias. Finally, by using methods that accept time-varying covariates, we avoided some of the oversimplifications and assumptions required in previous studies and accommodated temporal changes in SARS-CoV-2 infection status.

Potential study limitations included the inability, due to small numbers of some outcomes, to stratify analyses on multiple covariates simultaneously. Whereas the original virus strain and the alpha variant predominated before SARS-CoV-2 vaccines became available, most of the population had received at least one dose of vaccine by the time the omicron variant emerged,^24^ preventing us from disentangling the respective contributions of variant and vaccination. Furthermore, while our results were adjusted for age, sex, and degree of comorbidity, and stratified by age, virus variant, and vaccination status, we cannot rule out residual confounding by other factors. Finally, the Danish population is fairly ethnically homogeneous, with a relatively flat socioeconomic gradient, and our findings may not be generalizable to other populations.

## Conclusions

Our CVD results are reassuring, as they indicate that any long-term excess risks associated with SARS-CoV-2 infection are limited to a few outcomes (pulmonary embolism, venous embolism/thromboembolism, arrhythmias) and the potential increases in risk are small. Our KD results also suggest that any persistent increases in long-term risk associated with SARS-CoV-2 infection are minimal; however, these findings need to be confirmed in other populations. Most importantly, in a majority-vaccinated population, long-term CVD and KD risks do not appear to differ for omicron-infected and uninfected persons.

## Data Availability

The data used in this study were retrieved from Danish national registers and do not belong to the authors but to the Danish Health Data Authority. The authors are therefore not permitted to make the data publicly available. However, upon receipt of approval from the Danish Data Protection Agency, data from the registers are available by applying to the Danish Health Data Authority via their online request system.

