## Supplemental Material for "SARS-CoV-2 Infection and Long-Term Risk of Cardiovascular and Renal Morbidity"

**DEFINITIONS**

**Exclusion Diagnoses**

We excluded persons with International Classification of Diseases, version 10 [ICD-10], codes for congenital syndromes with defects in multiple organ systems (Q87.0-Q87.9 or Q89.3) or chromosomal abnormalities (Q90-Q99) registered in the Danish National Patient Register before the start of follow-up.

Additional exclusions, cardiovascular morbidity cohort

Cardiovascular disease: I00-I77, I79-I82, I87, I97, I98.0, I98.1, I98.8

Congenital heart defects: Q20-Q28

Other relevant heart-related diagnoses: B33.2 (viral carditis); C38.0, C38.8, or D15.1 (malignant or benign neoplasm of the heart); S25-S26 (injury of the heart or of the blood vessels of the thorax); Z94.1 or Z94.3 (heart transplant, with or without concomitant lung transplant); Z95.0 (cardiac pacemaker).

Additional exclusions, renal morbidity cohort

Renal disease: N00-N19, N25-N29, N99.0

Congenital kidney defects: Q27.1, Q27.2, Q60-Q63

Other relevant kidney-related diagnoses: A98.5 (hemorrhagic fever with renal syndrome); B52.0 (malaria with nephropathy); B90.1 (sequelae of genitourinary tuberculosis); C64-65, D30.0, D30.1, D41.0, or D41.1 (malignant, benign or uncertain neoplasm of kidney or renal pelvis); E10.2, E11.2, E13.2, or E14.2 (renal complications of type 1, type 2, other or unspecified diabetes mellitus); I13 (hypertensive heart and renal disease); I15.0 (renovascular hypertension); I15.1 (hypertension secondary to other renal disorders); S35.4 or S37.0 (injury of renal blood vessels or of kidney); Z94.0 (kidney transplant); Z99.2 (dependence on renal dialysis); BDJD0 or BDJD2 (renal dialysis, acute or chronic).

**Definition of Severe SARS-CoV-2 Infection**

We defined severe SARS-CoV-2 infection as a positive PCR test for SARS-CoV-2 less than 14 days before or within 2 days after a registered hospital admission or an outpatient contact lasting at least 12 hours, where a COVID-19-related ICD-10 code was registered in the National Patient Register in connection with the hospital contact. COVID-19-related ICD-10 codes included B34.2, B34.2A, B94.8A, B97.2, B972.A, B972.B, B97.2B1, and Z03.8PA1.

**Cardiovascular Disease Groups (Outcomes)**

We identified persons with incident cardiovascular disease during follow-up based on registration of the following groups of ICD-10 codes in the National Patient Register:

Ischemic heart disease: I20-25, with the exception of I25.2-25.4.

    Myocardial infarction only: I21

Pulmonary embolism: I26

Pericarditis and myocarditis: I30-31, I40

Valve disorders, including endocarditis: I33-38

Cardiomyopathy: I42.0-42.5, I42.8, I42.9

Heart failure: I50

Conduction disorders: I44-45

Arrhythmias: I47-49

Cardiac arrest: I46

Cerebrovascular hemorrhage: I60-62

Cerebral infarction: I63-64

Other cerebrovascular disease: I65-66, I67.2, I67.6, I67.8, I67.9, G45

Aneurysm/dissection (I25.3, I25.4, I67.0, I67.1, I71-72

Arterial embolism or thrombosis: I74

Venous embolism or thrombosis: I80-82.

**Kidney Disease Groups (Outcomes)**

We identified persons with incident renal disease during follow-up based on registration of the following groups of ICD-10 codes in the National Patient Register:

Acute kidney injury: N17

Other acute kidney disease: N00-01, N10
Chronic renal failure: N18
Other chronic kidney disease: N03, N11
Unspecified kidney disease involving renal failure: N19
Other unspecified kidney disease: N04-05, N12
Dialysis dependence without registration of an accompanying renal disease diagnosis: JFD0 (acute need for dialysis), JFD2 (chronic dialysis).

Supplementary Table 1. Hazard ratios for 15 cardiovascular disease groups comparing persons with a positive SARS-CoV-2 test and persons with only negative SARS-CoV-2 test results, by length of follow-up after the first positive test in the study period, Denmark, 1 March 2020-31 December 2022.

| Length of follow-up after first positive test* | | Ischemic heart disease | | | | Cerebral infarction | | | | Cerebrovascular hemorrhage | | | | Other cerebrovascular disease | | | |
|---|---|---|---|---|---|---|---|---|---|---|---|---|---|---|---|---|---|
| | Person-years | No. of events | Rate† | HR | 95% CI | No. of events | Rate† | HR | 95% CI | No. of events | Rate† | HR | 95% CI | No. of events | Rate† | HR | 95% CI |
| No positive test | 6,188,401 | 8415 | 1.4 | 1 | Ref | 6064 | 1.0 | 1 | Ref | 1503 | 0.2 | 1 | Ref | 4287 | 0.7 | 1 | Ref |
| Day 1 | 6814 | 36 | 5.3 | 5.84 | 4.21-8.11 | 36 | 5.3 | 8.29 | 5.97-11.5 | 8 | 1.2 | 6.93 | 3.45-13.9 | 26 | 3.8 | 8.75 | 5.95-12.9 |
| <1 month | 201,905 | 181 | 0.9 | 1.01 | 0.87-1.17 | 147 | 0.7 | 1.16 | 0.99-1.37 | 40 | 0.2 | 1.19 | 0.87-1.63 | 99 | 0.5 | 1.15 | 0.94-1.40 |
| 1-5 months | 1,101,947 | 950 | 0.9 | 0.93 | 0.87-1.00 | 638 | 0.6 | 0.88 | 0.81-0.96 | 164 | 0.1 | 0.86 | 0.73-1.01 | 501 | 0.5 | 1.01 | 0.92-1.11 |
| 6-11 months | 1,115,292 | 886 | 0.8 | 1.03 | 0.93-1.14 | 608 | 0.5 | 0.91 | 0.83-0.99 | 175 | 0.2 | 0.96 | 0.82-1.13 | 472 | 0.4 | 1.02 | 0.93-1.12 |
| ≥12 months | 295,266 | 274 | 0.9 | 0.89 | 0.78-1.00 | 135 | 0.5 | 0.74 | 0.62-0.88 | 46 | 0.2 | 0.85 | 0.63-1.14 | 108 | 0.4 | 0.77 | 0.64-0.94 |

| Length of follow-up after first positive test* | | Venous embolism/thrombosis | | | | Arterial embolism/thrombosis | | | | Pulmonary embolism | | | | Aneurysm or dissection | | | |
|---|---|---|---|---|---|---|---|---|---|---|---|---|---|---|---|---|---|
| | Person-years | No. of events | Rate† | HR | 95% CI | No. of events | Rate† | HR | 95% CI | No. of events | Rate† | HR | 95% CI | No. of events | Rate† | HR | 95% CI |
| Negative test | 6,188,401 | 4926 | 0.8 | 1 | Ref | 201 | 0.03 | 1 | Ref | 2425 | 0.4 | 1 | Ref | 2370 | 0.4 | 1 | Ref |
| Day 1 | 6814 | 11 | 1.6 | 2.67 | 1.48-4.84 | 0 | 0 | - | - | 27 | 4.0 | 16.1 | 11.0-23.5 | 7 | 1.0 | 3.85 | 1.83-8.09 |
| <1 month | 201,905 | 170 | 0.8 | 1.42 | 1.22-1.66 | 6 | 0.03 | 1.24 | 0.55-2.82 | 201 | 1.0 | 4.14 | 3.57-4.79 | 48 | 0.2 | 0.90 | 0.68-1.21 |
| 1-5 months | 1,101,947 | 751 | 0.7 | 1.12 | 1.03-1.21 | 21 | 0.02 | 0.80 | 0.50-1.26 | 337 | 0.3 | 1.21 | 1.08-1.36 | 306 | 0.3 | 1.02 | 0.90-1.15 |
| 6-11 months | 1,115,292 | 708 | 0.6 | 1.09 | 1.00-1.18 | 16 | 0.01 | 0.64 | 0.38-1.07 | 237 | 0.2 | 0.89 | 0.78-1.02 | 276 | 0.2 | 0.98 | 0.86-1.11 |
| ≥12 months | 295,266 | 180 | 0.6 | 0.90 | 0.77-1.05 | 3 | 0.01 | 0.39 | 0.12-1.24 | 73 | 0.2 | 0.77 | 0.61-0.98 | 57 | 0.2 | 0.73 | 0.56-0.95 |

| Length of follow-up after first positive test* | | Cardiac arrest | | | | Heart failure | | | | Cardiomyopathy | | | | Inflammatory heart disease | | | |
|---|---|---|---|---|---|---|---|---|---|---|---|---|---|---|---|---|---|
| | Person-years | No. of events | Rate† | HR | 95% CI | No. of events | Rate† | HR | 95% CI | No. of events | Rate† | HR | 95% CI | No. of events | Rate† | HR | 95% CI |
| Negative test | 6,188,401 | 833 | 0.1 | 1 | Ref | 1902 | 0.3 | 1 | Ref | 424 | 0.1 | 1 | Ref | 1037 | 0.2 | 1 | Ref |
| Day 1 | 6814 | 10 | 1.5 | 15.2 | 8.13-28.4 | 15 | 2.2 | 10.8 | 6.51-18.0 | 1 | 0.1 | 2.69 | 0.38-19.1 | 5 | 0.7 | 4.90 | 2.04-11.8 |
| <1 month | 201,905 | 23 | 0.1 | 1.21 | 0.80-1.84 | 40 | 0.2 | 1.00 | 0.73-1.37 | 9 | 0.04 | 0.83 | 0.42-1.60 | 32 | 0.2 | 1.08 | 0.76-1.54 |
| 1-5 months | 1,101,947 | 101 | 0.1 | 0.93 | 0.76-1.15 | 199 | 0.2 | 0.87 | 0.73-1.01 | 54 | 0.05 | 0.88 | 0.66-1.17 | 163 | 0.1 | 0.99 | 0.84-1.18 |
| 6-11 months | 1,115,292 | 75 | 0.1 | 0.73 | 0.57-0.92 | 157 | 0.1 | 0.73 | 0.62-0.87 | 66 | 0.06 | 1.13 | 0.86-1.47 | 175 | 0.2 | 1.05 | 0.89-1.23 |
| ≥12 months | 295,266 | 19 | 0.1 | 0.59 | 0.37-0.93 | 36 | 0.1 | 0.57 | 0.41-0.80 | 12 | 0.04 | 0.73 | 0.41-1.31 | 46 | 0.2 | 0.91 | 0.67-1.23 |

| | | Arrhythmias | | | | Conduction disorders | | | | Valve disorders |
|---|---|---|---|---|---|---|---|---|---|---|

| Length of follow-up after first positive test* | Person-years | No. of events | Rate† | HR | 95% CI | No. of events | Rate† | HR | 95% CI | No. of events | Rate† | HR | 95% CI |
|---|---|---|---|---|---|---|---|---|---|---|---|---|---|
| Negative test | 6,188,401 | 14,039 | 2.3 | 1 | Ref | 1769 | 0.3 | 1 | Ref | 4052 | 0.7 | 1 | Ref |
| Day 1 | 6814 | 82 | 12.0 | 7.67 | 6.17-9.53 | 12 | 1.8 | 8.85 | 5.01-15.6 | 7 | 1.0 | 2.51 | 1.20-5.28 |
| <1 month | 201,905 | 369 | 1.8 | 1.19 | 1.07-1.32 | 30 | 0.1 | 0.76 | 0.53-1.09 | 83 | 0.4 | 1.03 | 0.83-1.28 |
| 1-5 months | 1,101,947 | 1885 | 1.7 | 1.07 | 1.02-1.12 | 242 | 0.2 | 1.08 | 0.94-1.24 | 457 | 0.4 | 0.98 | 0.89-1.08 |
| 6-11 months | 1,115,292 | 1769 | 1.6 | 1.05 | 1.00-1.11 | 244 | 0.2 | 1.13 | 0.98-1.30 | 446 | 0.4 | 1.03 | 0.93-1.14 |
| ≥12 months | 295,266 | 433 | 1.5 | 0.90 | 0.82-0.99 | 69 | 0.2 | 1.13 | 0.88-1.44 | 107 | 0.4 | 0.86 | 0.71-1.04 |

CI, confidence interval. HR, hazard ratio.
* For most persons in whom a negative test preceded a later positive test (scenario B in Supplementary Figure 1), the "Day 1" and "<1 month" categories combined were synonymous with the acute phase of infection. However, in persons whose first test in the study period was positive (scenario A, Supplementary Figure 1) or whose positive test was returned within 30 days of a first, negative test in the study period, the "<1 month" category might stretch into the post-acute phase of infection (>30 days) due to the washout period.
† Rate per 1000 person-years, unadjusted.

Supplementary Table 2. Hazard ratios for 6 kidney disease groups comparing persons with a positive SARS-CoV-2 test and persons with only negative SARS-CoV-2 test results, by length of follow-up after the first positive test in the study period, Denmark, 1 March 2020-31 December 2022.

| Length of follow-up after first positive test* | Person-years | Acute kidney injury | | | | Other acute kidney disease | | | | Chronic renal failure | | | |
|---|---|---|---|---|---|---|---|---|---|---|---|---|---|
| | | No. of events | Rate† | HR | 95% CI | No. of events | Rate† | HR | 95% CI | No. of events | Rate† | HR | 95% CI |
| Negative test | 7,187,331 | 3591 | 0.5 | 1 | Ref | 3082 | 0.4 | 1 | Ref | 1807 | 0.3 | 1 | Ref |
| Day 0 | 7,521 | 63 | 8.4 | 28.6 | 22.2-36.7 | 20 | 2.7 | 6.96 | 4.48-10.8 | 6 | 0.8 | 4.81 | 2.15-10.7 |
| <1 month | 222,686 | 146 | 0.8 | 2.29 | 1.94-2.71 | 97 | 0.4 | 1.16 | 0.95-1.42 | 41 | 0.2 | 1.15 | 0.84-1.57 |
| 1-5 months | 1,215,287 | 305 | 0.3 | 0.83 | 0.74-0.94 | 420 | 0.3 | 0.86 | 0.78-0.96 | 177 | 0.1 | 0.88 | 0.75-1.03 |
| 6-11 months | 1,198,074 | 263 | 0.2 | 0.80 | 0.71-0.91 | 494 | 0.4 | 1.08 | 0.98-1.19 | 213 | 0.2 | 1.24 | 1.07-1.43 |
| ≥12 months | 315,507 | 79 | 0.3 | 0.76 | 0.60-0.95 | 130 | 0.4 | 1.08 | 0.91-1.30 | 44 | 0.1 | 1.13 | 0.83-1.54 |

| Length of follow-up after first positive test* | Person-years | Other chronic kidney disease | | | | Unspecified renal failure | | | | Other/unspecified kidney disease | | | |
|---|---|---|---|---|---|---|---|---|---|---|---|---|---|
| | | No. of events | Rate† | HR | 95% CI | No. of events | Rate† | HR | 95% CI | No. of events | Rate† | HR | 95% CI |
| Negative test | 7,187,331 | 125 | 0.02 | | | 756 | 0.1 | | | 2758 | 0.4 | | |
| Day 0 | 7,521 | 1 | 0.1 | 8.60 | 1.20-61.8 | 8 | 1.1 | 16.4 | 8.13-32.9 | 24 | 3.2 | 10.1 | 6.72-15.0 |
| <1 month | 222,686 | 2 | 0.01 | 0.60 | 0.15-2.44 | 15 | 0.1 | 1.05 | 0.63-1.76 | 95 | 0.4 | 1.36 | 1.11-1.68 |
| 1-5 months | 1,215,287 | 16 | 0.01 | 0.85 | 0.50-1.45 | 48 | 0.04 | 0.60 | 0.44-0.80 | 427 | 0.4 | 1.09 | 0.99-1.22 |
| 6-11 months | 1,198,074 | 23 | 0.02 | 1.29 | 0.82-2.05 | 85 | 0.1 | 1.22 | 0.97-1.54 | 469 | 0.4 | 1.22 | 1.10-1.35 |
| ≥12 months | 315,507 | 6 | 0.02 | 1.44 | 0.62-3.33 | 22 | 0.1 | 1.30 | 0.84-2.01 | 158 | 0.5 | 1.25 | 1.06-1.47 |

CI, confidence interval. HR, hazard ratio.

* For most persons in whom a negative test preceded a later positive test (scenario B in Supplementary Figure 1), the “Day 1” and “<1 month” categories combined were synonymous with the acute phase of infection. However, in persons whose first test in the study period was positive (scenario A, Supplementary Figure 1) or whose positive test was returned within 30 days of a first, negative test in the study period, the “<1 month” category might stretch into the post-acute phase of infection (>30 days) due to the washout period.

† Rate per 1000 person-years, unadjusted.

Supplementary Table 3. Hazard ratios for cardiovascular disease and kidney disease 1-12 months after the start of follow-up, comparing persons with a positive SARS-CoV-2 test and persons with only negative SARS-CoV-2 test results, by age at SARS-CoV-2 test, Denmark, 1 March 2020-31 December 2022.

| Outcome | Age at SARS-CoV-2 test | | | | | |
|---|---|---|---|---|---|---|
| | <40 years | | 40-64 years | | ≥65 years | |
| | HR | 95% CI | HR | 95% CI | HR | 95% CI |
| CARDIOVASCULAR DISEASE | | | | | | |
| Ischemic heart disease | 0.88 | 0.67-1.16 | 0.91 | 0.85-0.97 | 0.95 | 0.87-1.04 |
| Cerebral infarction | 0.93 | 0.72-1.20 | 0.89 | 0.82-0.98 | 0.89 | 0.81-0.97 |
| Cerebrovascular hemorrhage | 0.88 | 0.65-1.19 | 0.97 | 0.82-1.14 | 0.82 | 0.66-1.01 |
| Other cerebrovascular disease | 0.96 | 0.72-1.27 | 1.03 | 0.93-1.14 | 1.02 | 0.92-1.13 |
| Venous embolism/thrombosis | 1.10 | 0.98-1.25 | 1.09 | 1.00-1.19 | 1.10 | 0.97-1.24 |
| Arterial embolism/thrombosis | 0.47 | 0.16-1.38 | 0.89 | 0.56-1.39 | 0.52 | 0.25-1.08 |
| Pulmonary embolism | 1.25 | 0.98-1.59 | 1.07 | 0.93-1.23 | 0.90 | 0.77-1.04 |
| Aneurysm/dissection | 1.16 | 0.84-1.59 | 1.05 | 0.92-1.19 | 0.91 | 0.78-1.06 |
| Cardiac arrest | 0.83 | 0.58-1.18 | 0.83 | 0.65-1.05 | 0.83 | 0.61-1.12 |
| Heart failure | 0.72 | 0.46-1.13 | 0.89 | 0.76-1.05 | 0.72 | 0.61-0.87 |
| Cardiomyopathy | 1.04 | 0.69-1.59 | 1.12 | 0.84-1.49 | 0.72 | 0.46-1.12 |
| Inflammatory heart disease | 1.00 | 0.84-1.18 | 1.13 | 0.92-1.39 | 0.78 | 0.49-1.23 |
| Arrhythmias | 1.06 | 0.97-1.15 | 1.09 | 1.03-1.15 | 1.02 | 0.96-1.09 |
| Conduction disorders | 1.22 | 1.00-1.48 | 0.96 | 0.80-1.15 | 1.16 | 0.98-1.36 |
| Valve disorders | 0.81 | 0.65-1.02 | 1.02 | 0.90-1.15 | 1.05 | 0.95-1.16 |
| KIDNEY DISEASE | | | | | | |
| Acute kidney injury | 0.98 | 0.73-1.31 | 0.74 | 0.62-0.89 | 0.81 | 0.72-0.91 |
| Other acute kidney disease | 0.97 | 0.88-1.07 | 0.87 | 0.73-1.03 | 1.10 | 0.91-1.33 |
| Chronic renal failure | 3.13 | 1.93-5.10 | 1.20 | 0.93-1.54 | 0.94 | 0.82-1.07 |
| Other chronic kidney disease | 1.07 | 0.62-1.85 | 1.04 | 0.54-2.03 | 1.05 | 0.49-2.26 |
| Unspecified renal failure | 3.07 | 1.47-6.40 | 1.07 | 0.72-1.60 | 0.75 | 0.60-0.95 |
| Other/unspecified kidney disease | 1.13 | 1.01-1.25 | 1.08 | 0.92-1.25 | 1.34 | 1.13-1.59 |

CI, confidence interval. HR, hazard ratio.

Supplementary Table 4. Hazard ratios for cardiovascular disease and kidney disease 1-12 months after the start of follow-up, comparing persons with a positive SARS-CoV-2 test and persons with only negative SARS-CoV-2 test results, excluding persons hospitalized for COVID-19, Denmark, 1 March 2020-31 December 2022.

| | Hazard Ratio | 95% Confidence Interval |
|---|---|---|
| CARDIOVASCULAR DISEASE | | |
| Ischemic heart disease | 0.93 | 0.88-0.98 |
| Cerebral infarction | 0.89 | 0.83-0.94 |
| Cerebrovascular hemorrhage | 0.89 | 0.79-1.01 |
| Other cerebrovascular disease | 1.02 | 0.95-1.10 |
| Venous embolism/thrombosis | 1.09 | 1.02-1.16 |
| Arterial embolism/thrombosis | 0.70 | 0.48-1.01 |
| Pulmonary embolism | 0.97 | 0.88-1.07 |
| Aneurysm/dissection | 0.99 | 0.90-1.09 |
| Cardiac arrest | 0.81 | 0.68-0.96 |
| Heart failure | 0.79 | 0.70-0.89 |
| Cardiomyopathy | 0.99 | 0.80-1.22 |
| Inflammatory heart disease | 1.02 | 0.89-1.16 |
| Arrhythmias | 1.05 | 1.01-1.09 |
| Conduction disorders | 1.09 | 0.98-1.21 |
| Valve disorders | 1.01 | 0.94-1.09 |
| KIDNEY DISEASE | | |
| Acute kidney injury | 0.76 | 0.69-0.84 |
| Other acute kidney disease | 0.97 | 0.90-1.05 |
| Chronic renal failure | 1.06 | 0.94-1.20 |
| Other chronic kidney disease | 1.06 | 0.72-1.54 |
| Unspecified renal failure | 0.90 | 0.74-1.10 |
| Other/unspecified kidney disease | 1.13 | 1.04-1.22 |

Supplementary Table 5. Hazard ratios for cardiovascular disease and kidney disease 1-12 months after the start of follow-up, comparing persons with a positive SARS-CoV-2 test and persons with only negative SARS-CoV-2 test results, by SARS-CoV-2 variant, Denmark, 1 March 2020-31 December 2022.

| | Presumed SARS-CoV-2 strain | | | | | | | |
|---|---|---|---|---|---|---|---|---|
| Outcome | Original strain | | Alpha | | Delta | | Omicron | |
| | HR | 95% CI | HR | 95% CI | HR | 95% CI | HR | 95% CI |
| CARDIOVASCULAR DISEASE | | | | | | | | |
| Ischemic heart disease | 0.86 | 0.72-1.02 | 0.94 | 0.69-1.26 | 0.89 | 0.69-1.15 | 0.95 | 0.89-1.01 |
| Cerebral infarction | 0.99 | 0.79-1.24 | 0.91 | 0.60-1.37 | 0.99 | 0.74-1.33 | 0.88 | 0.82-0.94 |
| Cerebrovascular hemorrhage | 0.90 | 0.59-1.38 | 0.85 | 0.40-1.79 | 1.24 | 0.77-2.00 | 0.92 | 0.79-1.05 |
| Other cerebrovascular disease | 1.09 | 0.86-1.39 | 0.86 | 0.54-1.39 | 1.03 | 0.74-1.44 | 1.02 | 0.94-1.11 |
| Venous embolism/thrombosis | 1.06 | 0.86-1.30 | 1.22 | 0.89-1.66 | 0.99 | 0.75-1.31 | 1.08 | 1.00-1.16 |
| Arterial embolism/thrombosis | 2.50 | 1.20-5.22 | 0.88 | 0.12-6.33 | 1.16 | 0.29-4.70 | 0.60 | 0.39-0.94 |
| Pulmonary embolism | 1.51 | 1.17-1.96 | 1.69 | 1.11-2.59 | 1.43 | 0.98-2.09 | 0.89 | 0.79-0.99 |
| Aneurysm/dissection | 1.08 | 0.79-1.50 | 1.04 | 0.59-1.84 | 0.79 | 0.48-1.32 | 1.04 | 0.93-1.15 |
| Cardiac arrest | 0.52 | 0.26-1.06 | 0.89 | 0.37-2.15 | 0.65 | 0.27-1.56 | 0.90 | 0.75-1.09 |
| Heart failure | 0.61 | 0.38-0.97 | 0.46 | 0.17-1.22 | 1.30 | 0.81-2.07 | 0.81 | 0.70-0.92 |
| Cardiomyopathy | 0.64 | 0.26-1.57 | 1.80 | 0.74-4.38 | 1.00 | 0.37-2.70 | 1.03 | 0.81-1.31 |
| Inflammatory heart disease | 1.52 | 1.06-2.17 | 1.46 | 0.87-2.44 | 1.22 | 0.77-1.92 | 0.92 | 0.78-1.07 |
| Arrhythmias | 1.10 | 0.96-1.25 | 0.92 | 0.73-1.17 | 1.19 | 1.01-1.40 | 1.04 | 1.00-1.09 |
| Conduction disorders | 0.79 | 0.52-1.21 | 1.38 | 0.82-2.35 | 0.66 | 0.38-1.17 | 1.18 | 1.04-1.33 |
| Valve disorders | 0.89 | 0.68-1.18 | 1.12 | 0.71-1.76 | 1.10 | 0.79-1.54 | 1.01 | 0.93-1.10 |
| KIDNEY DISEASE | | | | | | | | |
| Acute kidney injury | 0.73 | 0.53-1.02 | 0.62 | 0.29-1.30 | 0.69 | 0.42-1.13 | 0.81 | 0.73-0.90 |
| Other acute kidney disease | 0.98 | 0.75-1.28 | 0.59 | 0.36-0.98 | 0.69 | 0.48-1.00 | 0.98 | 0.90-1.08 |
| Chronic renal failure | 0.45 | 0.24-0.88 | 0.82 | 0.26-2.55 | 0.77 | 0.36-1.61 | 1.09 | 0.96-1.23 |
| Other chronic kidney disease | - | - | 2.51 | 0.61-10.3 | 1.34 | 0.33-5.47 | 1.04 | 0.67-1.60 |
| Unspecified renal failure | 0.36 | 0.12-1.13 | 1.25 | 0.31-5.04 | 1.18 | 0.49-2.85 | 0.85 | 0.68-1.05 |
| Other/unspecified kidney disease | 1.38 | 1.10-1.74 | 1.62 | 1.18-2.23 | 1.36 | 1.02-1.80 | 1.02 | 0.93-1.13 |

CI, confidence interval. HR, hazard ratio.

Supplementary Table 6. Hazard ratios for cardiovascular disease and kidney disease 1-12 months after the start of follow-up, comparing persons with a positive SARS-CoV-2 test and persons with only negative SARS-CoV-2 test results, by vaccination status at SARS-CoV-2 test, Denmark, 1 March 2020-31 December 2022.

| Outcome | Number of vaccine doses received at the time of SARS-CoV-2 testing | | | | | | | |
|---|---|---|---|---|---|---|---|---|
| | None | | One | | Two | | Three or more | |
| | HR | 95% CI | HR | 95% CI | HR | 95% CI | HR | 95% CI |
| CARDIOVASCULAR DISEASE | | | | | | | | |
| Ischemic heart disease | 0.89 | 0.79-1.00 | 0.86 | 0.53-1.38 | 0.90 | 0.77-1.05 | 0.94 | 0.85-1.04 |
| Cerebral infarction | 1.05 | 0.91-1.22 | 0.77 | 0.43-1.38 | 0.89 | 0.74-1.08 | 0.86 | 0.76-0.98 |
| Cerebrovascular hemorrhage | 1.09 | 0.85-1.39 | 1.10 | 0.44-2.80 | 0.80 | 0.56-1.12 | 0.87 | 0.68-1.11 |
| Other cerebrovascular disease | 0.97 | 0.82-1.16 | 0.99 | 0.53-1.83 | 0.88 | 0.71-1.10 | 1.07 | 0.92-1.23 |
| Venous embolism/thrombosis | 1.11 | 0.98-1.26 | 0.98 | 0.64-1.49 | 1.09 | 0.92-1.28 | 1.10 | 0.96-1.25 |
| Arterial embolism/thrombosis | 1.24 | 0.65-2.37 | - | - | 0.54 | 0.17-1.67 | 0.61 | 0.30-1.23 |
| Pulmonary embolism | 1.44 | 1.20-1.72 | 1.29 | 0.67-2.50 | 1.12 | 0.85-1.46 | 0.86 | 0.71-1.04 |
| Aneurysm/dissection | 0.98 | 0.78-1.22 | 0.54 | 0.17-1.73 | 0.94 | 0.71-1.25 | 1.03 | 0.85-1.24 |
| Cardiac arrest | 0.75 | 0.53-1.07 | 1.06 | 0.37-3.04 | 0.80 | 0.50-1.27 | 0.86 | 0.62-1.19 |
| Heart failure | 0.84 | 0.64-1.10 | 0.44 | 0.11-1.80 | 0.89 | 0.63-1.25 | 0.78 | 0.62-0.97 |
| Cardiomyopathy | 1.24 | 0.83-1.85 | - | - | 0.60 | 0.31-1.16 | 1.04 | 0.67-1.62 |
| Inflammatory heart disease | 1.26 | 1.00-1.58 | 1.83 | 1.08-3.10 | 0.93 | 0.69-1.27 | 0.86 | 0.64-1.16 |
| Arrhythmias | 1.00 | 0.92-1.09 | 1.07 | 0.80-1.43 | 1.11 | 1.00-1.23 | 1.06 | 0.98-1.15 |
| Conduction disorders | 1.10 | 0.87-1.37 | 0.55 | 0.22-1.37 | 0.95 | 0.71-1.25 | 1.20 | 0.95-1.50 |
| Valve disorders | 0.84 | 0.70-1.00 | 0.99 | 0.53-1.83 | 1.09 | 0.88-1.35 | 1.04 | 0.89-1.21 |
| KIDNEY DISEASE | | | | | | | | |
| Acute kidney injury | 0.98 | 0.80-1.20 | 1.62 | 0.88-2.98 | 0.94 | 0.72-1.23 | 0.73 | 0.61-0.87 |
| Other acute kidney disease | 0.94 | 0.84-1.06 | 1.05 | 0.68-1.62 | 0.96 | 0.79-1.17 | 1.01 | 0.83-1.23 |
| Chronic renal failure | 0.73 | 0.50-1.06 | 1.78 | 0.69-4.61 | 2.48 | 1.76-3.49 | 0.95 | 0.75-1.21 |
| Other chronic kidney disease | 0.31 | 0.10-1.01 | 0.89 | 0.10-7.91 | 1.49 | 0.60-3.70 | 1.35 | 0.59-3.13 |
| Unspecified renal failure | 0.76 | 0.43-1.34 | 4.21 | 1.34-13.2 | 1.47 | 0.83-2.58 | 0.80 | 0.55-1.18 |
| Other/unspecified kidney disease | 1.33 | 1.17-1.52 | 1.13 | 0.72-1.76 | 1.15 | 0.95-1.39 | 1.01 | 0.85-1.21 |

CI, confidence interval. HR, hazard ratio.

Supplementary Table 7. Hazard ratios for 15 cardiovascular disease groups comparing persons with a positive SARS-CoV-2 test and persons with only negative SARS-CoV-2 test results, by length of follow-up after the first positive test in the study period, when follow-up ended on 10 March 2022.

| Length of follow-up after first positive test* | Person-years | Ischemic heart disease | | | Cerebral infarction | | | Cerebrovascular hemorrhage | | | Other cerebrovascular disease | | | Venous embolism or thrombosis | | |
|---|---|---|---|---|---|---|---|---|---|---|---|---|---|---|---|---|
| | | No. of events | HR | 95% CI | No. of events | HR | 95% CI | No. of events | HR | 95% CI | No. of events | HR | 95% CI | No. of events | HR | 95% CI |
| No positive test | 4,732,238 | 6110 | 1 | Ref | 4100 | 1 | Ref | 1035 | 1 | Ref | 3107 | 1 | Ref | 3622 | 1 | Ref |
| Day 1 | 6161 | 12 | 2.44 | 1.39-4.31 | 17 | 5.38 | 3.33-8.68 | 5 | 5.58 | 2.43-14.22 | 15 | 6.35 | 3.81-10.58 | 5 | 1.54 | 0.64-3.70 |
| <1 month | 163,013 | 108 | 0.92 | 0.76-1.12 | 93 | 1.25 | 1.02-1.55 | 25 | 1.19 | 0.80-1.79 | 69 | 1.24 | 0.97-1.59 | 122 | 1.51 | 1.26-1.81 |
| 1-5 months | 242,882 | 183 | 0.92 | 0.80-1.07 | 116 | 0.98 | 0.81-1.18 | 29 | 0.82 | 0.57-1.19 | 97 | 1.05 | 0.86-1.29 | 159 | 1.16 | 0.99-1.37 |
| 6-11 months | 118,442 | 104 | 0.83 | 0.68-1.00 | 57 | 0.80 | 0.62-1.05 | 16 | 0.77 | 0.47-1.27 | 46 | 0.81 | 0.60-1.09 | 75 | 0.92 | 0.73-1.16 |
| ≥12 months | 50,715 | 66 | 1.06 | 0.83-1.35 | 21 | 0.60 | 0.39-0.92 | 5 | 0.50 | 0.21-1.21 | 19 | 0.67 | 0.43-1.06 | 34 | 0.89 | 0.63-1.25 |

| Length of follow-up after first positive test* | Person-years | Arterial embolism or thrombosis | | | Pulmonary embolism | | | Aneurysm or dissection | | | Cardiac arrest | | | Heart failure | | |
|---|---|---|---|---|---|---|---|---|---|---|---|---|---|---|---|---|
| | | No. of events | HR | 95% CI | No. of events | HR | 95% CI | No. of events | HR | 95% CI | No. of events | HR | 95% CI | No. of events | HR | 95% CI |
| Negative test | 4,732,238 | 137 | 1 | Ref | 1677 | 1 | Ref | 1655 | 1 | Ref | 570 | 1 | Ref | 1264 | 1 | Ref |
| Day 1 | 6161 | 0 | - | - | 12 | 10.4 | 5.88-18.5 | 0 | - | - | 4 | 7.99 | 2.97-21.5 | 6 | 5.97 | 2.66-13.4 |
| <1 month | 163,013 | 3 | 0.93 | 0.29-2.99 | 162 | 5.84 | 4.91-6.84 | 30 | 0.88 | 0.61-1.26 | 14 | 1.15 | 0.67-1.94 | 22 | 0.95 | 0.62-1.46 |
| 1-5 months | 242,882 | 8 | 1.70 | 0.83-3.49 | 119 | 2.15 | 1.79-2.60 | 69 | 1.30 | 1.02-1.66 | 18 | 0.88 | 0.55-1.40 | 39 | 1.00 | 0.73-1.38 |
| 6-11 months | 118,442 | 4 | 1.51 | 0.55-4.15 | 25 | 0.66 | 0.44-0.98 | 20 | 0.63 | 0.40-0.98 | 9 | 0.72 | 0.37-1.49 | 13 | 0.52 | 0.30-0.90 |
| ≥12 months | 50,715 | 2 | 1.56 | 0.38-6.38 | 14 | 0.75 | 0.44-1.27 | 8 | 0.51 | 0.25-1.02 | 3 | 0.51 | 0.16-1.59 | 9 | 0.73 | 0.38-1.41 |

| Length of follow-up after first positive test* | Person-years | Cardiomyopathy | | | Inflammatory heart disease | | | Arrhythmias | | | Conduction disorders | | | Valve disorders | | |
|---|---|---|---|---|---|---|---|---|---|---|---|---|---|---|---|---|
| | | No. of events | HR | 95% CI | No. of events | HR | 95% CI | No. of events | HR | 95% CI | No. of events | HR | 95% CI | No. of events | HR | 95% CI |
| Negative test | 4,732,238 | 295 | 1 | Ref | 804 | 1 | Ref | 9931 | 1 | Ref | 1295 | 1 | Ref | 2817 | 1 | Ref |
| Day 1 | 6161 | 0 | - | - | 2 | 2.05 | 0.51-8.24 | 41 | 4.82 | 3.55-6.56 | 2 | 1.81 | 0.45-7.25 | 5 | 2.32 | 0.96-5.60 |
| <1 month | 163,013 | 8 | 1.13 | 0.55-2.31 | 22 | 0.88 | 0.57-1.36 | 229 | 1.11 | 0.97-1.27 | 20 | 0.74 | 0.47-1.15 | 45 | 0.89 | 0.66-1.20 |
| 1-5 months | 242,882 | 8 | 0.71 | 0.35-1.43 | 51 | 1.30 | 0.98-1.73 | 386 | 1.16 | 1.05-1.29 | 40 | 0.89 | 0.65-1.22 | 99 | 1.19 | 0.97-1.45 |
| 6-11 months | 118,442 | 8 | 1.21 | 0.59-2.46 | 30 | 1.50 | 1.04-2.17 | 198 | 1.02 | 0.88-1.18 | 23 | 0.91 | 0.60-1.38 | 38 | 0.75 | 0.54-1.03 |
| ≥12 months | 50,715 | 1 | 0.32 | 0.05-2.30 | 6 | 0.71 | 0.32-1.58 | 94 | 1.00 | 0.82-1.23 | 13 | 1.10 | 0.64-1.91 | 22 | 0.87 | 0.57-1.32 |

CI, confidence interval. HR, hazard ratio.

* For most persons in whom a negative test preceded a later positive test (scenario B in Supplementary Figure 1), the “Day 1” and “<1 month” categories combined were synonymous with the acute phase of infection. However, in persons whose first test in the study period was positive (scenario A, Supplementary Figure 1) or whose positive test was returned within 30 days of a first, negative test in the study period, the “<1 month” category might stretch into the post-acute phase of infection (>30 days) due to the washout period.

Supplementary Table 8. Hazard ratios for 6 kidney disease groups comparing persons with a positive SARS-CoV-2 test and persons with only negative SARS-CoV-2 test results, by length of follow-up after the first positive test in the study period, when follow-up ended on 10 March 2022.

| Length of follow-up after first positive test* | Person-years | Acute kidney injury | | | Other acute kidney disease | | | Chronic renal failure | | |
|---|---|---|---|---|---|---|---|---|---|---|
| | | No. of events | HR | 95% CI | No. of events | HR | 95% CI | No. of events | HR | 95% CI |
| Negative test | 5,435,056 | 2270 | 1 | Ref | 2335 | 1 | Ref | 892 | 1 | Ref |
| Day 0 | 6,688 | 24 | 17.8 | 11.9-26.8 | 12 | 4.83 | 2.74-8.54 | 0 | - | - |
| <1 month | 175,496 | 93 | 3.01 | 2.43-3.72 | 73 | 1.14 | 0.90-1.44 | 15 | 1.48 | 0.88-2.49 |
| 1-5 months | 261,750 | 73 | 1.23 | 0.97-1.55 | 93 | 0.85 | 0.69-1.04 | 15 | 0.72 | 0.43-1.19 |
| 6-11 months | 129,925 | 31 | 0.75 | 0.53-1.08 | 54 | 1.01 | 0.77-1.33 | 11 | 0.70 | 0.39-1.98 |
| ≥12 months | 56,601 | 18 | 0.83 | 0.52-1.32 | 23 | 1.03 | 0.68-1.55 | 4 | 0.47 | 0.17-1.25 |

| Length of follow-up after first positive test* | Person-years | Other chronic kidney disease | | | Unspecified renal failure | | | Other/unspecified kidney disease | | |
|---|---|---|---|---|---|---|---|---|---|---|
| | | No. of events | HR | 95% CI | No. of events | HR | 95% CI | No. of events | HR | 95% CI |
| Negative test | 7,187,331 | 90 | | | 415 | | | 2138 | | |
| Day 0 | 7,521 | 0 | - | - | 5 | 26.2 | 10.7-64.2 | 13 | 6.09 | 3.52-10.5 |
| <1 month | 222,686 | 2 | 0.94 | 0.23-3.89 | 5 | 1.15 | 0.47-2.81 | 75 | 1.37 | 1.08-1.74 |
| 1-5 months | 1,215,287 | 2 | 0.54 | 0.13-2.19 | 3 | 0.32 | 0.10-1.00 | 140 | 1.48 | 1.24-1.76 |
| 6-11 months | 1,198,074 | 1 | 0.50 | 0.07-3.60 | 6 | 0.88 | 0.39-1.98 | 94 | 1.79 | 1.45-2.21 |
| ≥12 months | 315,507 | 0 | - | - | 3 | 0.81 | 0.26-2.53 | 30 | 1.29 | 0.90-1.85 |

CI, confidence interval. HR, hazard ratio.

* For most persons in whom a negative test preceded a later positive test (scenario B in Supplementary Figure 1), the “Day 1” and “<1 month” categories combined were synonymous with the acute phase of infection. However, in persons whose first test in the study period was positive (scenario A, Supplementary Figure 1) or whose positive test was returned within 30 days of a first, negative test in the study period, the “<1 month” category might stretch into the post-acute phase of infection (>30 days) due to the washout period.

Supplementary Figure 1. Illustration of the way in which follow-up time was accumulated according to SARS-CoV-2 test status

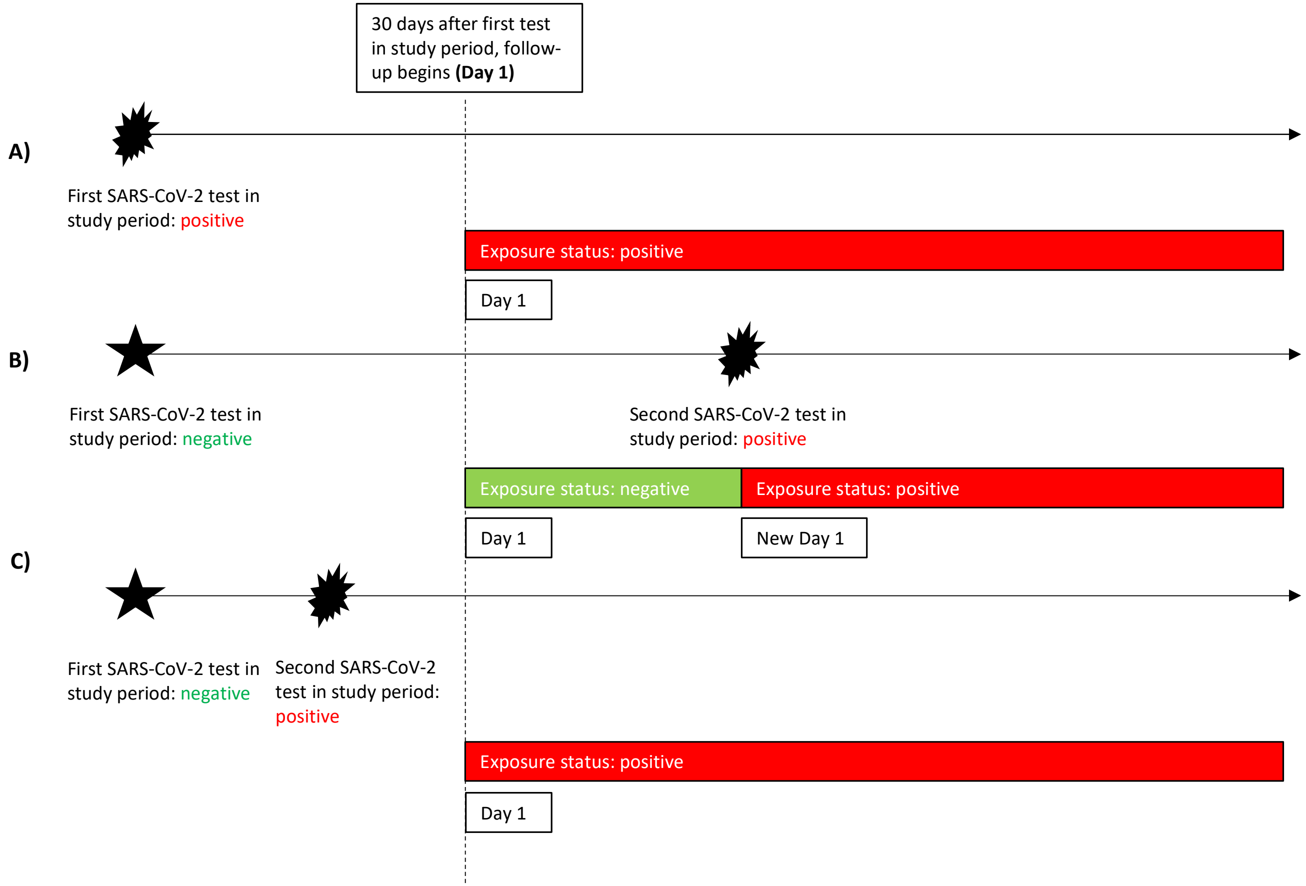

In scenario A), the first SARS-CoV-2 test in the study period was positive. This person entered follow-up 30 days later as test-positive at Day 1 and contributed follow-up time to the test-postive group until the study ended.

In scenario B), the first SARS-CoV-2 test in the study period was negative. This person entered follow-up 30 days later as test-negative at Day 1 and contributed follow-up time to the test-negative group until s/he received a second test and was found to be infected. On the date of the second test, exposure status changed to test-positive and the day counter reverted to Day 1. This person then contributed follow-up time to the test-positive group until the study ended.

In scenario C), the first SARS-CoV-2 test in the study period was negative. This person then had a second, positive SARS-CoV-2 test before the original 30-day washout period ended. Consequently, this person entered follow-up 30 days after the first (negative) test as test-positive at Day 1 and contributed follow-up time to the test-positive group until the study ended.

In all three cases, once follow-up began after a positive test, all follow-up time was considered test-positive, regardless of any subsequent negative tests. Furthermore, the day counter did not revert to Day 1 at any subsequent positive tests.

Supplementary Figure 2. Distribution of follow-up time across the study period by exposure status. At each point in time, the figure shows the percentage of the study cohort contributing time in each exposure group (green, uninfected, SARS-CoV-2 test negative; orange, infected, ≤30 days from a positive SARS-CoV-2 test]; blue, infected, >30 days from a positive SARS-CoV-2 test). Note that at the start of the study period, very few persons had been tested for SARS-CoV-2 infection; as a result, in the first 3 months of the study, a small number of infected individuals represented a large proportion of the persons contributing follow-up, producing an initial spike that faded as the number of tests increased dramatically.

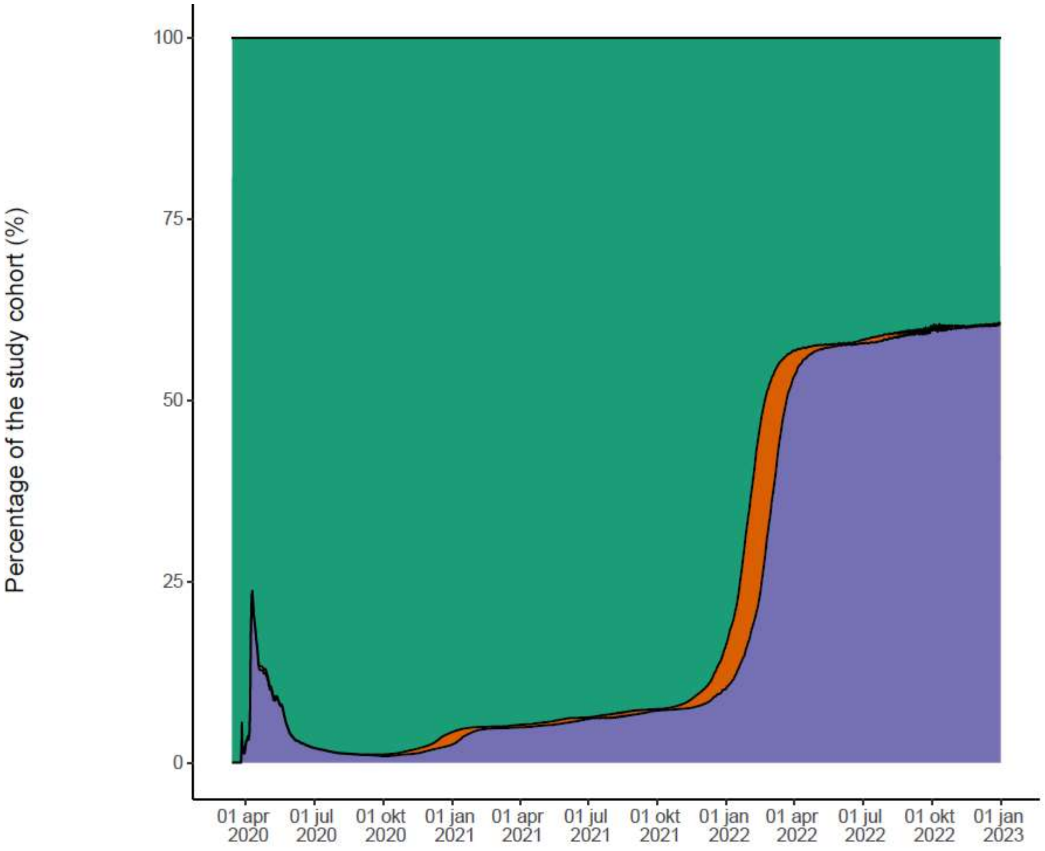

Study Period

| Individuals in cohort | | | | | | | | | | | | |
|---|---|---|---|---|---|---|---|---|---|---|---|---|
| Uninfected | 350 | 383.4K | 1.4M | 2.7M | 3.3M | 3.6M | 3.7M | 3.4M | 1.8M | 1.8M | 1.7M | 1.7M |
| Infected, day 1-30 | 1 | 126 | 3.5K | 48.2K | 12.2K | 8K | 8.8K | 250.7K | 149.2K | 20.9K | 12K | 11.1[illegible] |
| Infected, >30 days | 8 | 7.9K | 12.4K | 70.1K | 172.7K | 232.2K | 286.8K | 423.2K | 2.3M | 2.5M | 2.6M | 2.6M |

Supplementary Figure 3. Hazard ratios for 15 cardiovascular disease outcomes and 6 kidney disease outcomes by SARS-CoV-2 infection status, stratified by age at SARS-CoV-2 test, in a cohort of 4,508,489 persons (cardiovascular disease) or 5,150,480 (kidney disease) in Denmark, 2020-2022. Hazard ratios compare hazards in the period 1-12 months after a positive SARS-CoV-2 test with those in persons who only had negative SARS-CoV-2 test results. Red, <40 years; green, 40-64 years; blue, ≥65 years.

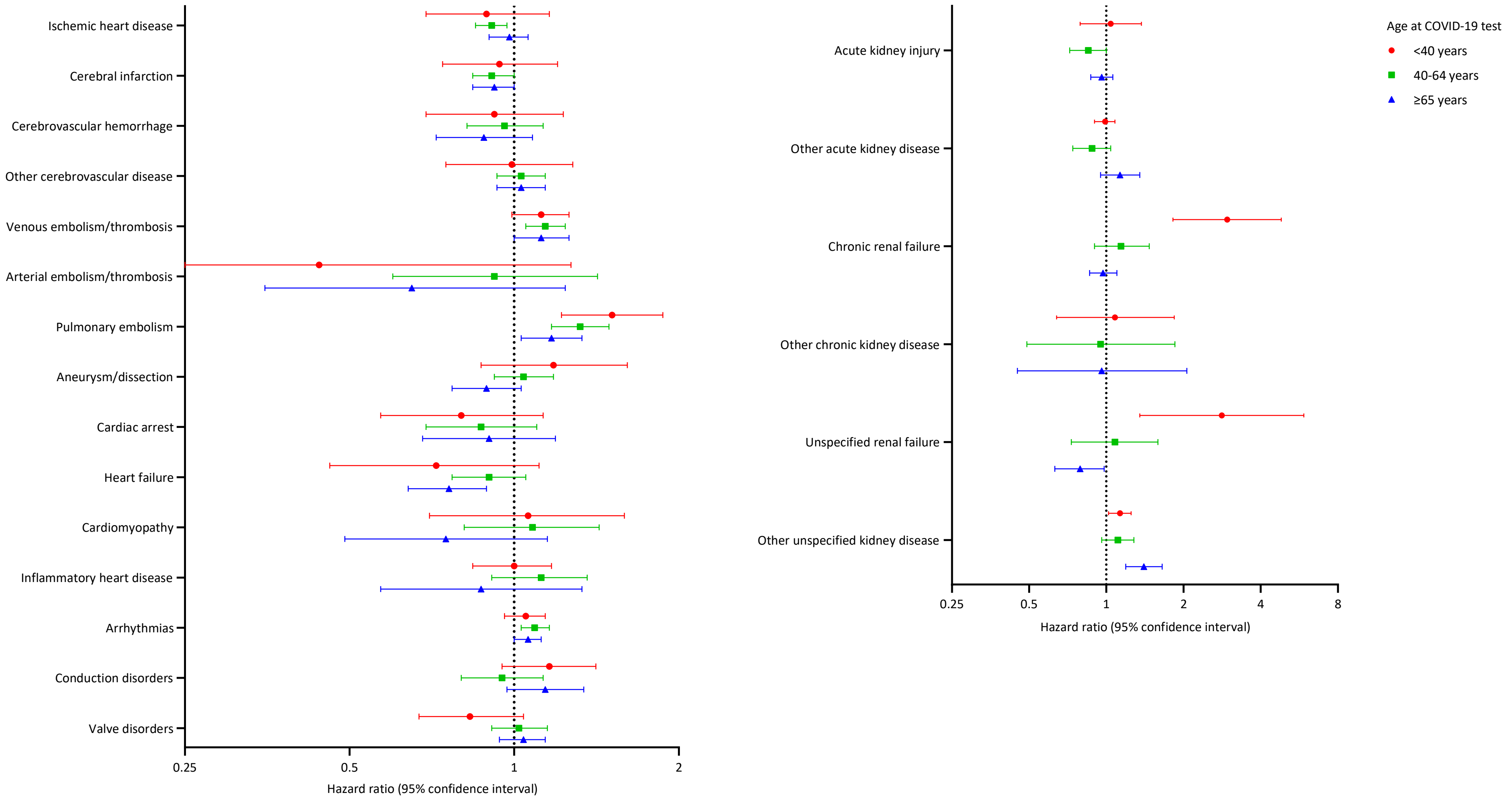

Supplementary Figure 4. Hazard ratios for 15 cardiovascular disease outcomes and 6 kidney disease outcomes by SARS-CoV-2 infection status, stratified by presumed SARS-CoV-2 variant, in a cohort of 4,508,489 persons (cardiovascular disease) or 5,150,480 (kidney disease) in Denmark, 2020-2022. Hazard ratios compare hazards in the period 1-12 months after a positive SARS-CoV-2 test with those in persons who only had negative SARS-CoV-2 test results.  Red, original SARS-CoV-2 virus; orange, alpha variant; green, delta variant; blue, omicron variant.

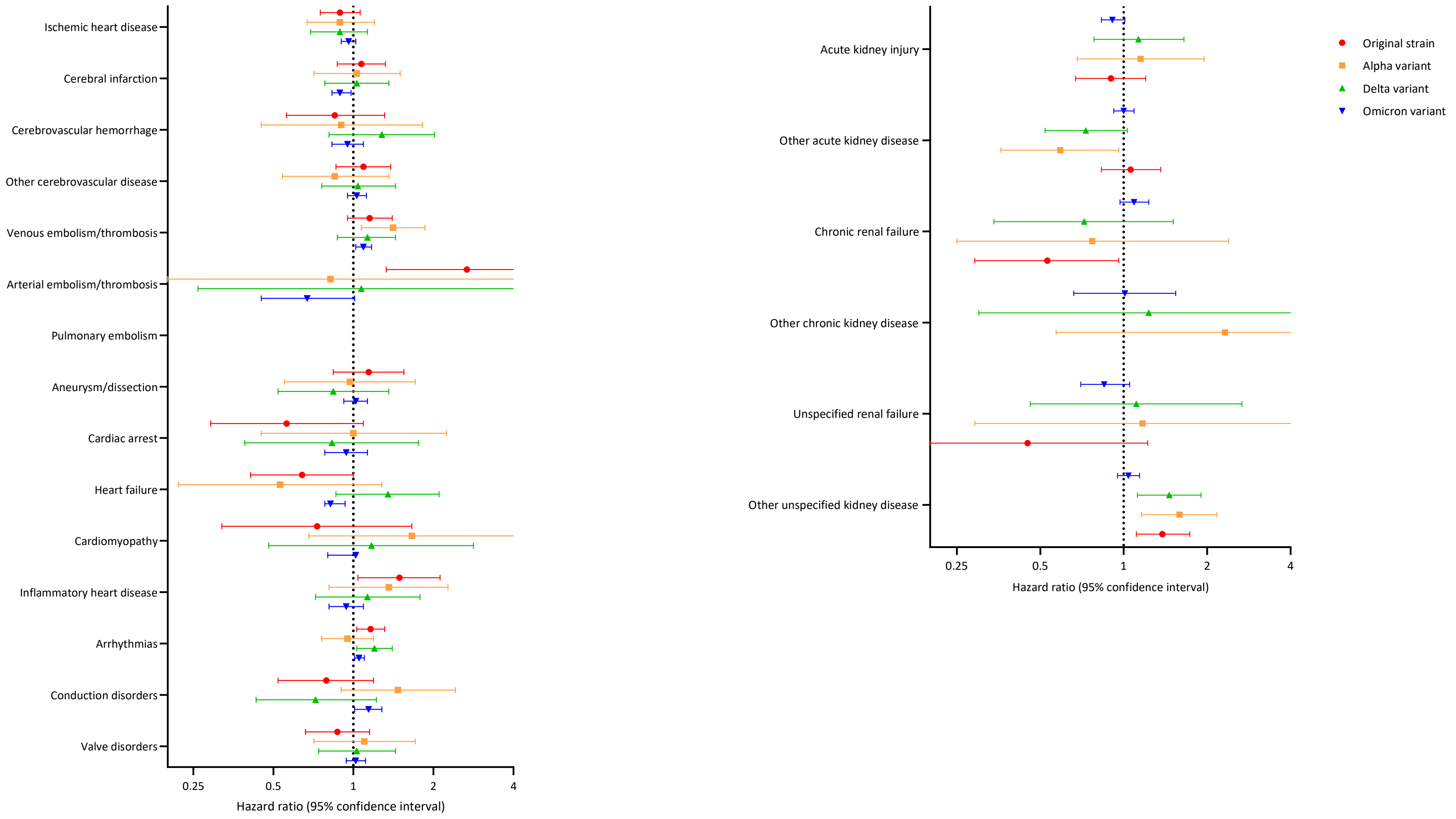

Supplementary Figure 5. Hazard ratios for 15 cardiovascular disease outcomes and 6 kidney disease outcomes by SARS-CoV-2 infection status, stratified by number of vaccine doses received before the SARS-CoV-2 test, in a cohort of 4,508,489 persons (cardiovascular disease) or 5,150,480 (kidney disease) in Denmark, 2020-2022. Hazard ratios compare hazards in the period 1-12 months after a positive SARS-CoV-2 test with those in persons who only had negative SARS-CoV-2 test results. Red, unvaccinated; orange, 1 dose; green, 2 doses; blue, ≥3 doses.

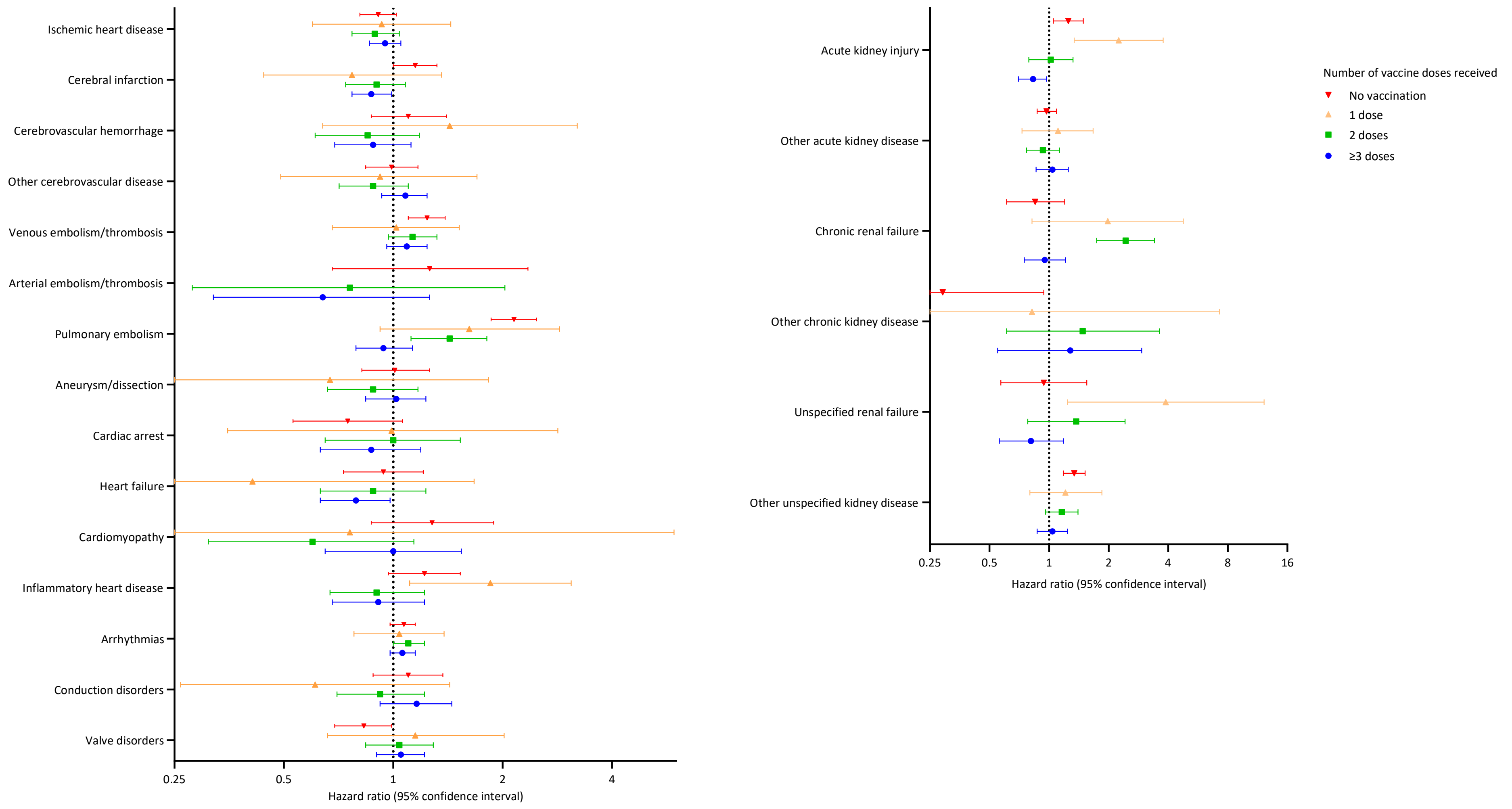
